# A Comparative Study of Alcohol Use, Alcohol Use Disorder and Consequences Among Young People and Adults with Injuries in the Northern Zone of Tanzania

**DOI:** 10.1101/2025.06.10.25329384

**Authors:** Winfrida C. Mwita, Elizabeth M. Keating, Rune Nathaniel Philemon, Baraka Moshi, Alice Andongolile, Florida J. Muro, Blandina T. Mmbaga, João Ricardo Nickenig Vissoci, Sia E. Msuya, Catherine A. Staton

## Abstract

**Background:** Alcohol use is a major risk factor for injuries, which are the leading cause of significant morbidity and mortality among young people in sub-Saharan Africa (SSA). In Tanzania, high rates of alcohol use disorders (AUD) and heavy episodic drinking have been documented. However, limited studies have examined age differences in alcohol use and related harm among injury patients. Understanding these differences is essential for designing targeted interventions, particularly for populations at heightened risk.

**Methods:** We conducted a cross-sectional analysis using linked data from the Kilimanjaro Christian Medical Centre (KCMC) Trauma Registry and the Pragmatic Randomized Adaptive Clinical Trial (PRACT). The sample included injured patients aged ≥18 years presenting within 24 hours of injury. Alcohol use was assessed using the Alcohol Use Disorders Identification Test (AUDIT), and alcohol-related consequences were measured using the Drinker Inventory of Consequences (DrInC). We compared prevalence of alcohol use disorder, risk levels, and consequences between young people (18–24 years) and adults (≥25 years) using descriptive statistics and age-stratified analyses.

**Results:** Of the 2,427 injury patients included, young people (18–24 years) had a mean age of 21.5 (SD 1.9), and adults (25+ years) had a mean age of 41.7 (SD 14.3). Of these, 46.5% of young people and 59.4% of adults reported being current drinkers. A higher proportion of adults had consumed alcohol in the past four weeks (28.7%) and within six hours before injury (21.3%) compared to young people (18.9% and 1.4%, respectively). Drinking frequency differed significantly, with adults more likely to drink four or more times per week than young people (14.1% vs. 5.8%, *p* < 0.001). Young people were significantly more likely to report that they or someone else had been injured as a result of their drinking within the past year (14.0% vs. 9.4%, *p* = 0.026). The majority of participants experienced alcohol-related consequences, with young people reporting significantly higher physical consequences (85.7%) than adults (73.5%) (*p* = 0.010). Despite these differences, the prevalence of alcohol use disorder (AUD) was comparable between the two groups (37.2% in young people vs. 39.6% in adults; *p* = 0.524).

**Conclusion:** Although adults reported more frequent alcohol consumption, young people experienced higher occurrences of alcohol-related harms, including a higher frequency of reporting that they or someone else had been injured due to their drinking, as well as having physical consequences. Despite these differences, both groups exhibited a comparable burden of problematic drinking, as measured by screening positive for AUD on the AUDIT. These findings highlight the need for age-specific interventions: for young people, strategies should target episodic, high-risk drinking behaviors associated with acute harms such as injury, while for adults, interventions may focus on habitual patterns of alcohol use.

## Introduction

Alcohol use remains a global public health crisis due to its substantial contribution to morbidity, mortality, and social harm. The World Health Organization (WHO) estimates that harmful alcohol use was responsible for 2.6 million deaths globally in 2019 and contributed to 115.9 million disability-adjusted life years (DALYs), with injuries accounting for over 27.6% of alcohol-attributable deaths (1). The burden is particularly pronounced in low- and middle-income countries (LMICs), especially in sub-Saharan Africa (SSA), where the age-standardized rate of alcohol-attributable deaths is among the highest globally (1). Young people (ages 15–24) are disproportionately affected, with alcohol being the leading contributor to injury, violence, and early mortality in this age group (1,2).

In Tanzania, this burden is particularly concerning. Previous studies have documented high rates of alcohol consumption and alcohol use disorders (AUD), especially among young men. For example, Francis et al. (2015) found that 10.5% of individuals aged 15–24 in Kilimanjaro and Mwanza regions of Tanzania screened positive for AUD (3), while Mbatia et al. (2009) reported a 5.7% prevalence of hazardous drinking among adults in Dar es Salaam (4). In clinical settings, alcohol use is also prominent among injury patients: a study conducted at a referral hospital in Kilimanjaro found that 30% of acutely injured patients tested positive for alcohol via breathalyzer on arrival to the emergency department (5). Despite this high burden, there is still limited research examining how alcohol-related risks and patterns differ by age, particularly among populations presenting with injuries. Injury patients represent a particularly relevant group for studying alcohol-related harm, as alcohol is a major proximal risk factor for both unintentional and intentional injuries, and emergency department settings offer a unique opportunity to assess these patterns across age groups (6,7).

Understanding how alcohol use behaviors and associated consequences differ across age groups is important for designing effective interventions. Studies have shown that drinking patterns may vary by age, shaped by social, cultural, and developmental factors—with younger individuals more likely to engage in episodic heavy drinking and adults tending toward more routine, habitual consumption (2,8,9). These differences are not only behavioral but also neurobiological: exposure to alcohol during adolescence and young adulthood—a period of ongoing brain development, can alter neural pathways related to impulse control and reward processing (10–12). Such changes heighten vulnerability to hazardous drinking patterns and increase the risk of developing AUD in the long term (13). Furthermore, early initiation of alcohol use has been linked to genetic predisposition for addiction and worsened mental health outcomes (14). In Northern Tanzania, Staton et al. (2020) found that individuals who initiated alcohol use before age 18 had significantly higher scores on both the AUDIT and DrInC scales, highlighting early drinking as a predictor of alcohol-related problems later in life (15).

These insights underscore the importance of examining age-specific drinking patterns, particularly among high-risk populations such as injury patients. While literature has documented age-related differences in alcohol consumption and its consequences, limited studies in Tanzania have compared these patterns specifically among injury patients. Some recent Tanzanian studies have begun to explore specific subgroups of injury patients. For instance, Pauley et al. (2023) documented notable gender differences in alcohol use patterns among emergency department patients, emphasizing the need for tailored interventions (16). However, much of the existing research continues to aggregate findings across age groups or focuses broadly on alcohol-related harms in the general population. This study addresses this gap by comparing alcohol use behaviors and alcohol-related consequences between young people and adults with injuries in the Northern zone of Tanzania. Findings are intended to inform age-appropriate public health interventions for reducing alcohol-related harms in this high-risk population.

## Methodology

### Study Design and Setting

This cross-sectional study was conducted at Kilimanjaro Christian Medical Center (KCMC), a tertiary referral hospital located in Moshi, in the Kilimanjaro region of the Northern zone of Tanzania. Tanzania ranks third in alcohol consumption across Africa (1), with the Kilimanjaro region reporting the highest and rapidly increasing levels of alcohol use in the country. KCMC serves as the regional referral hospital for the Northern Zone, and its Emergency Department (ED) manages approximately 2,000 injury cases annually (17) . Ethical approval to conduct this study was obtained from the Kilimanjaro Christian Medical University College Ethics Committee (Certificate #2708) and the Tanzanian National Institute of Medical Research (NIMR/HQ/R.8a/Vol.IX/4839).

### Data Source

This analysis included two linked data sources: the KCMC Trauma Registry and the KCMC Pragmatic Randomized Adaptive Clinical Trial (PRACT) Registry. The Trauma Registry prospectively collects routine clinical and demographic data for all patients aged 18 years and older who present to the ED with acute injuries. It includes information on demographics, injury type and mechanism, vital signs, clinical care, and ED outcomes. Patients are excluded if they present for follow-up care or if they die in the ED before evaluation.

The PRACT trial, a randomized controlled trial evaluating a brief alcohol intervention among injury patients, provides additional research-specific data. This includes self-reported alcohol use prior to injury, Alcohol Use Disorders Identification Test (AUDIT) scores, breathalyzer results (BAC > 0.0 g/dL), and Drinker Inventory of Consequences (DrInC) scores. Data were collected only after participants were clinically sober and provided informed consent. PRACT received ethical approvals from Duke University Medical Center IRB (Pro000103724), the KCMUCo Ethics Committee (Certificate #2457), and the National Institute for Medical Research (NIMR/HQ/R.8a/Vol.IX/3425). Further methodological details of PRACT are published elsewhere (18). The trial is registered at ClinicalTrials.gov: NCT04535011. Data used for this study were collected between October 2020 and February 2025 and were accessed for research purposes in March 2025, following ethical approval.

### Study Population

This study included injury patients aged 18 years and above who presented at KCMC’s ED within 24 hours of injury occurrence. Only patients who were clinically sober and provided informed consent were included. For analysis purposes, patients were divided into two groups: (1) young people (18 to 24 years) and (2) adults (25 years and above).

### Measures

Sociodemographic information, including age, sex, tribe, religion, highest educational attainment, employment status, and monthly income, was obtained from the Trauma Registry. Alcohol use data was obtained from the PRACT trial, which collects detailed information on participants’ alcohol consumption using structured questionnaires and the Timeline Followback (TLFB) method, a calendar-based, interviewer-administered tool designed to enhance recall of substance use over a specified time period (19). This study examined the prevalence of alcohol use across various time points: within the past 12 months, the past 28 days, and 6 hours prior to injury. Alcohol Use Disorder was assessed using the WHO Alcohol Use Disorders Identification Test (AUDIT) among current drinkers, defined as those who had consumed a drink containing alcohol in the last 12 months (20), with a score of ≥8 indicating AUD (21). The AUDIT tool has been validated for use among injury patients in Tanzania (22). Patterns of alcohol use were further explored among patients who reported alcohol consumption within the month prior to the injury. These patterns included frequency and quantity of alcohol consumed, with one standard drink defined as containing 10 g of pure ethanol (23). The DrInC questionnaire was used to assess the adverse consequences of alcohol abuse (16), also administered as part of the PRACT trial. DrInC measures five categories: interpersonal, physical, social, impulsive, and intrapersonal aspects. The physical subscale captures adverse physical states resulting from excessive drinking, such as hangovers, sleep disturbances, vomiting, health deterioration, changes in appearance, eating problems, sexual problems, and injuries while drinking. The interpersonal subscale assesses harms to relationships, including conflicts with family and friends, damaged social life, and loss of close relationships. The social responsibility subscale refers to the impact of drinking on fulfilling expected roles, including missing work or school, financial problems, and legal troubles. The impulse control subscale involves risky or impulsive behaviors related to drinking, such as fights, driving under the influence, injuries to oneself or others, and property damage. Lastly, the intrapersonal subscale measures internal emotional and psychological impacts, including feelings of guilt, shame, unhappiness, personality changes, and hindered personal growth. Each category employs a timeframe focusing on the past 3 months as well as a lifetime measure of alcohol consequences. For the present study, recent (past 3 months) alcohol-related adverse consequences have been considered.

### Statistical Analysis

Data management and analysis were conducted using STATA version 15 statistical software (17). Descriptive statistics were used to summarize participant characteristics, alcohol use patterns, and alcohol-related consequences, stratified by age group (young people aged 18–24 years and adults aged 25 years and older). Categorical variables were summarized using frequencies and proportions, while continuous variables were assessed for normality and summarized using means with standard deviations or medians with interquartile ranges (IQR), as appropriate. Comparisons between young people and adults were conducted using Pearson’s chi-square tests for categorical variables. Fisher’s exact tests were applied where expected cell counts were less than five. For continuous variables, independent t-tests were used for normally distributed data, while the Mann-Whitney U test was applied for non-normally distributed variables. Alcohol use prevalence was examined across different timeframes (past year, past four weeks, and within six hours before injury). Drinking risk levels and patterns were categorized based on the AUDIT tool, while alcohol-related consequences were assessed using the DrInC subscales, with comparisons between age groups. Statistical significance was set at *p* < 0.05 for all comparisons.

## Results

### Socio-demographic characteristics of the study participants

A total of 2,427 injury patients were included in the study, with 520 young people (18-24 years) and 1,907 adults (25+ years). The mean age of the participants was 37.3 years (SD ± 15.1). The majority of participants were male (80.4%), with young people showing a significantly higher proportion of males (86.0%) compared to adults (78.8%). Regarding the tribe, most participants were Chagga (43.7%), followed by Pare (13.2%), with significant differences between the age groups in tribal distribution. In terms of education, young people had a significantly higher proportion of secondary education completion (46.6%) compared to adults (22.6%), while the proportions of participants with vocational or college/university education were similar between the two age groups. Employment status revealed that while most participants were employed (92.8%), young people had a significantly higher proportion of being students (15.1%) compared to adults (0.6%). Additionally, young people earned a significantly lower median monthly income of TZS 150,000 (IQR: 50,000–300,000) compared to adults, who earned a median of TZS 240,000 (IQR: 100,000–450,000), (Table **1**)

**Table 1:**
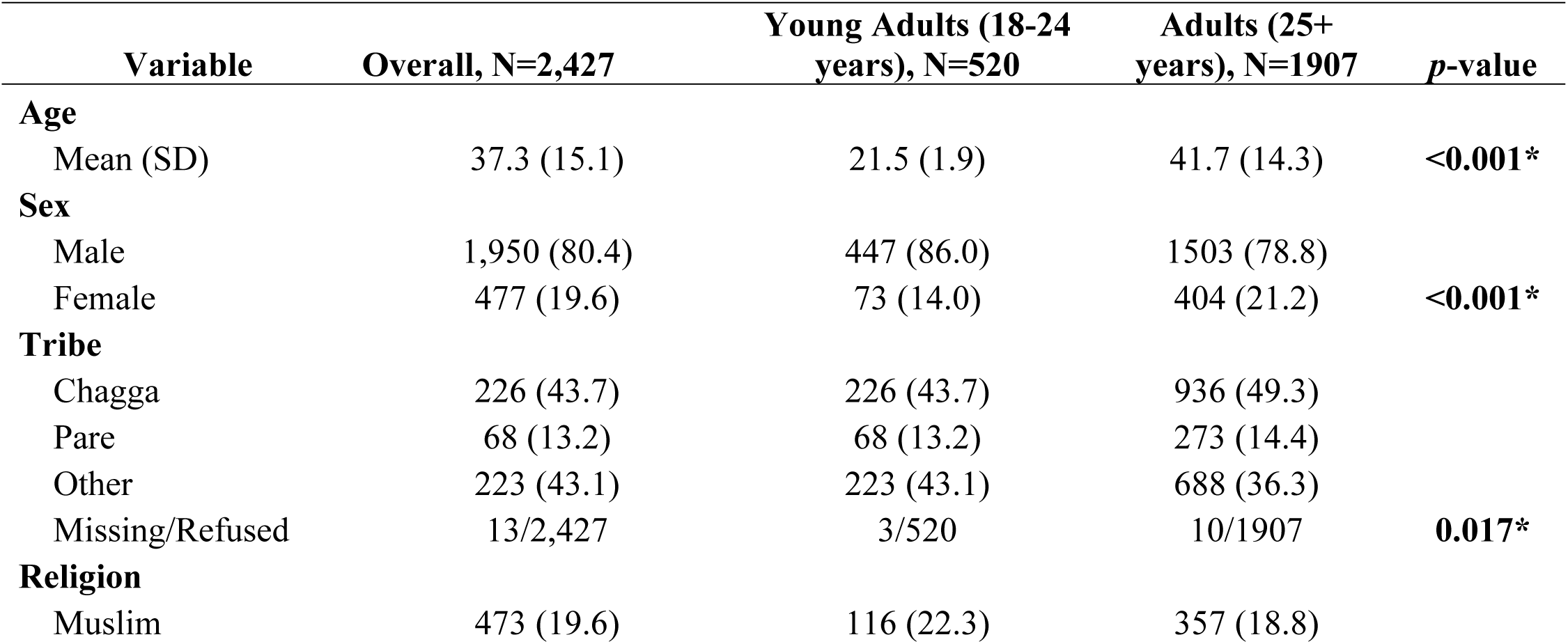
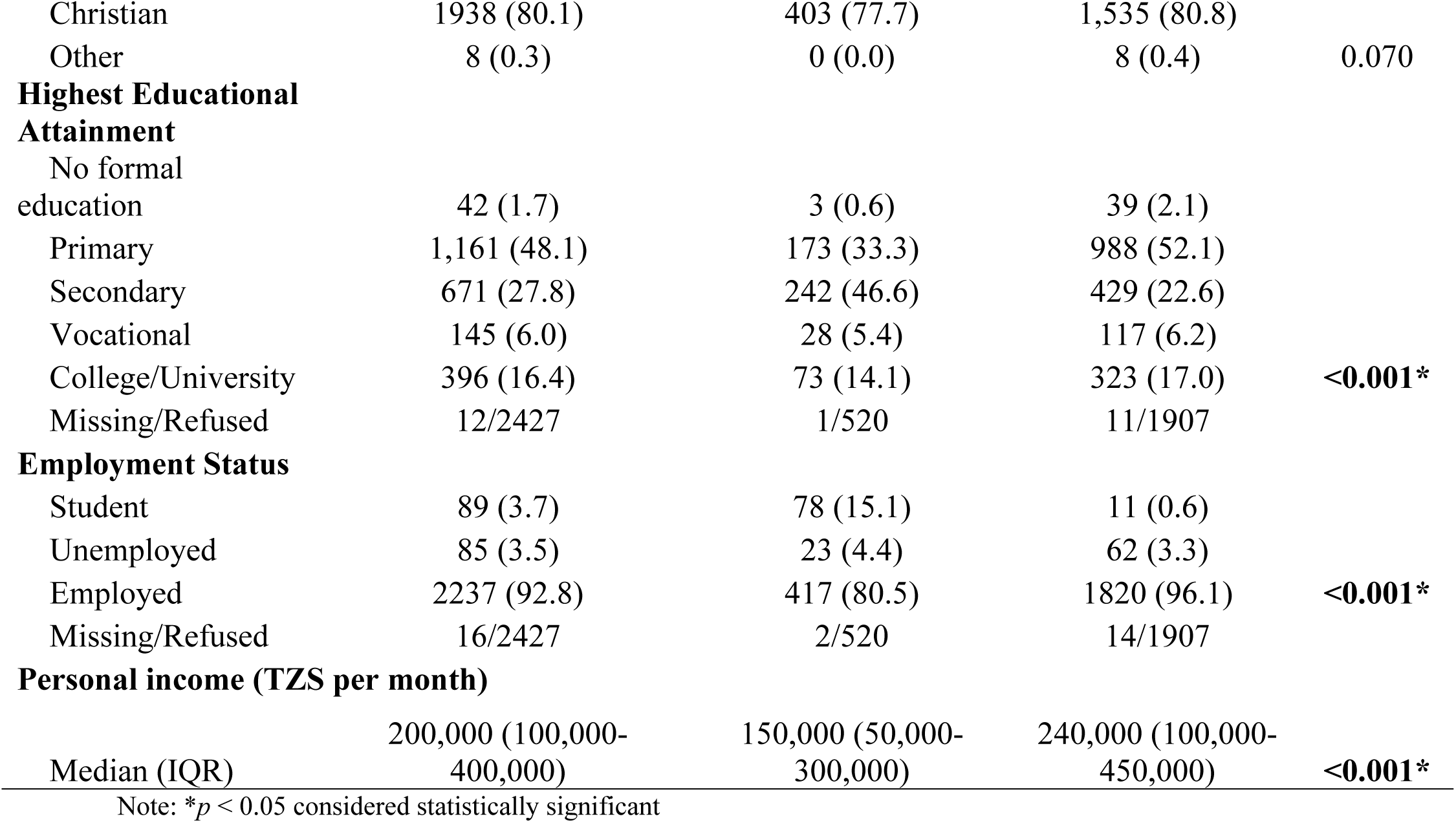
Socio-Demographic Characteristics of Injury Patients in the Northern Zone of Tanzania by Age Group (N=2,427)

### Prevalence of alcohol use among Young Adults and Adult injury patients in Northern Tanzania

Figure 1 shows the reported alcohol use across three timeframes. Adults consistently reported higher levels of alcohol consumption across all timeframes compared to young people. Specifically, 59.4% (1,130/1,903) of adults vs. 46.5% (242/520) of young people were current drinkers, having consumed alcohol in the past 12 months; 28.7% (547/1,907) of adults vs. 18.9% (98/520) consumed alcohol in the last four weeks; and 21.3% (405/1,905) of adults vs. 11.4% (59/520) reported alcohol use within six hours before injury.

**Figure 1:**
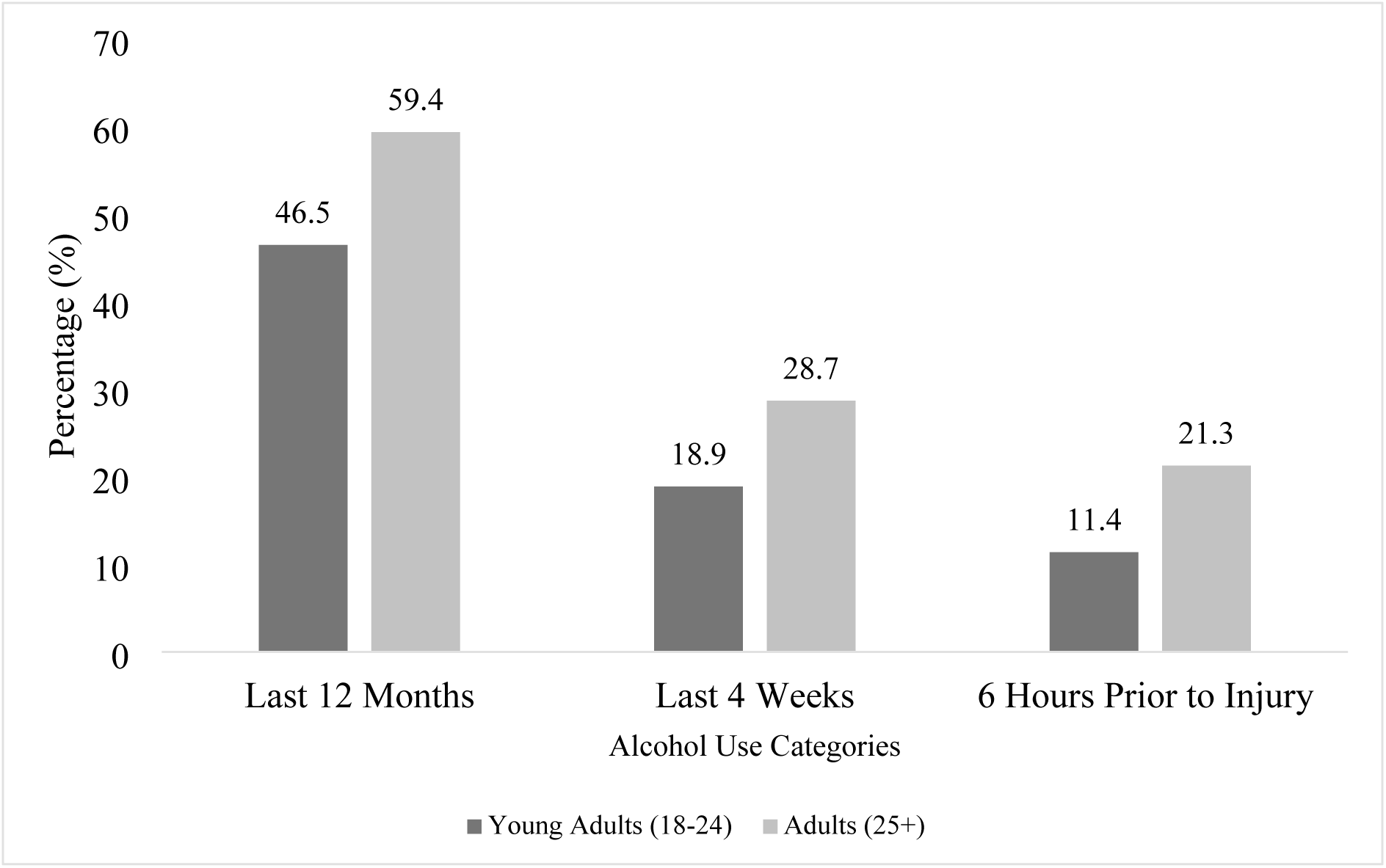
Reported Alcohol Use Among Injury Patients by Age Group - Young People (18-24, n= 520) vs Adults (25+, n = 1,907)

### Drinking Risk Levels and AUD among Young People Versus Adults

Table **2** presents the distribution of AUDIT drinking risk levels among current drinkers, defined as participants who reported alcohol use within the past 12 months. The majority of participants (60.8%) fell into the low-risk category (AUDIT score 1–7), with 62.8% of young people and 60.4% of adults in this group. The proportions in other risk categories—AUDIT scores of 8–15, 16–19, and 20–40 were also similar between age groups. Although adults had a slightly higher proportion in the highest risk category (9.3%) compared to young people (7.4%), none of the differences across individual categories were statistically significant. Additionally, 37.2% of young people and 39.6% of adults met criteria for alcohol use disorder (AUD), defined as an AUDIT score of 8 or higher. This difference was also not statistically significant (*p* = 0.524).

**Table 2:** AUDIT Drinking Risk Levels and AUD Prevalence by Age Group Among Current Drinkers (N = 1,372)

| AUDIT Drinking Risk Level | Total | Young People, n=242) | Adults, n=1,130 | *p*-value |
| --- | --- | --- | --- | --- |
| AUDIT Score 1-7 | 834 (60.8) | 152 (62.8) | 682 (60.4) |  |
| AUDIT Score 8-15 | 324 (23.6) | 56 (23.1) | 268 (23.7) |  |
| AUDIT Score 16-19 | 91 (6.6) | 16 (6.6) | 75 (6.6) |  |
| AUDIT Score 20-40 | 123 (9.0) | 18 (7.4) | 105 (9.3) |  |
| *Overall difference across AUDIT scores* |  |  |  | 0.804 |
| Total AUDIT ≥8 (AUD) | 538 (39.2%) | 90 (37.2%) | 448 (39.6%) | 0.524 |

Note: \* $p < 0.05$ considered statistically significant

### Drinking Pattern among Injury Patients

Table **3** presents the drinking pattern among injury patients who are current drinkers. A significant difference was observed in the frequency of alcohol consumption (*p* < 0.001). A higher proportion of young people (46.7%) reported drinking alcohol monthly or less compared to 34.3% of adults, while adults were more likely to consume alcohol four or more times per week (14.1%) compared to young people (5.8%). In addition, a significant difference was found in self-reported injuries to oneself or others due to one’s own drinking (*p* = 0.026), with 14.0% of young people and 9.4% of adults reporting that they or someone else had been injured as a result of their drinking in the past year.

**Table 3:**
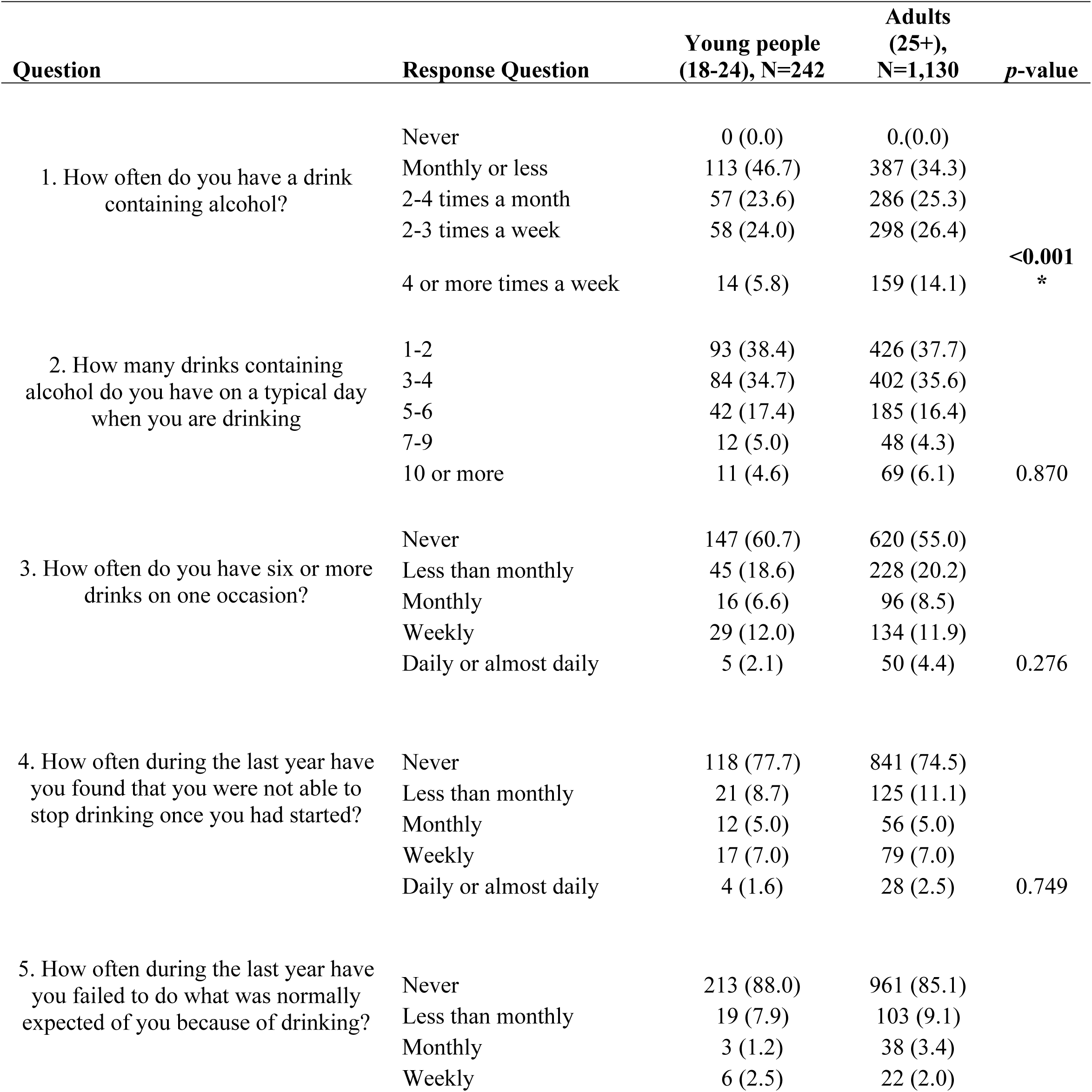
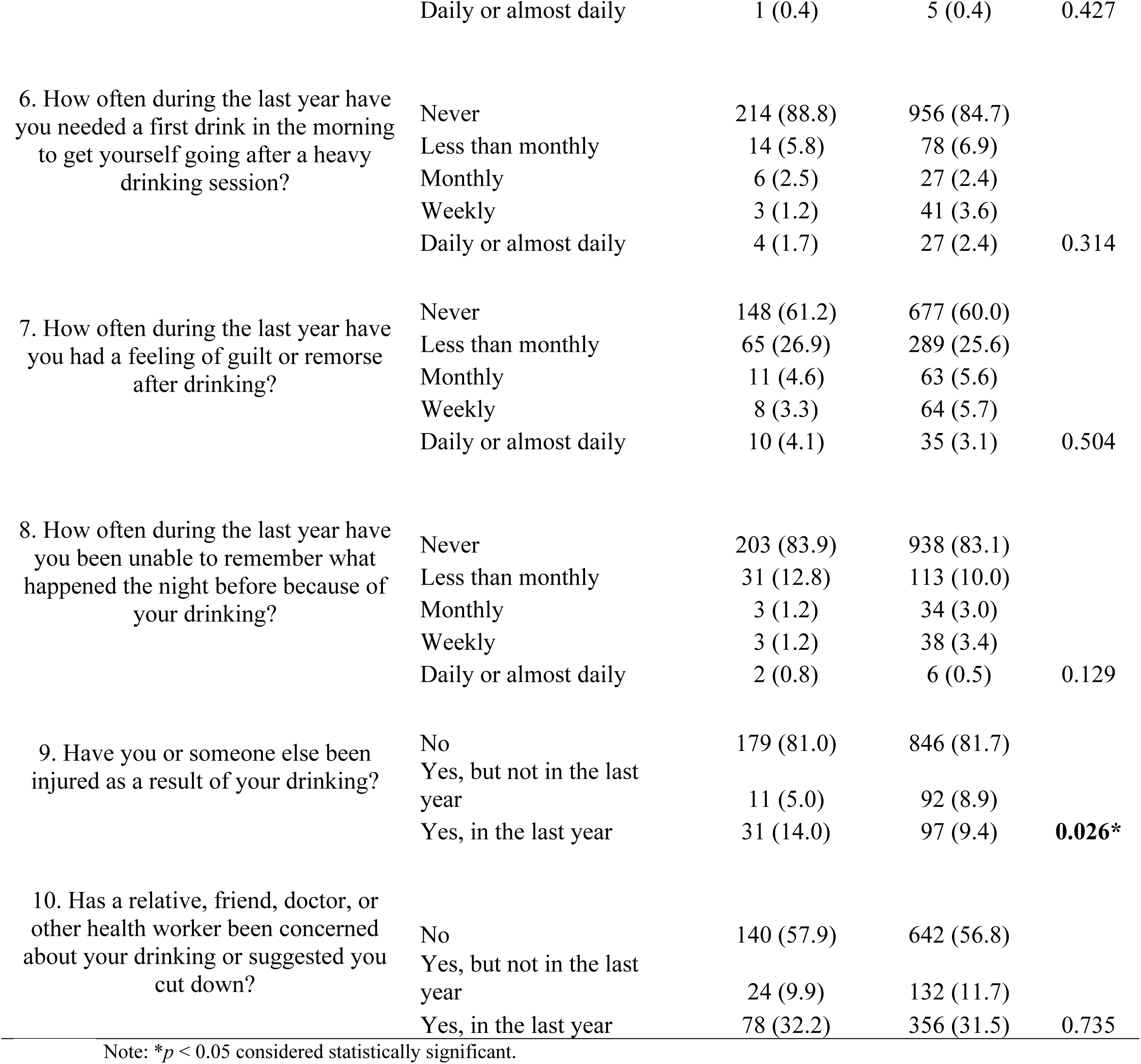
Pattern of Drinking Behavior among Injury Patients: Comparison between Young people and Adults (N = 1,372)

While the frequency of alcohol use and self-reported injury due to one’s own drinking differed significantly by age group, the remaining AUDIT-based responses showed no significant differences. These included the typical number of drinks consumed per occasion and the frequency of binge drinking episodes. Responses related to potential alcohol dependence, such as difficulty stopping drinking once started, needing a morning drink to recover from heavy drinking, and failing to meet responsibilities due to alcohol use—were also comparable between young people and adults. Similarly, alcohol-related harms, including feelings of guilt or remorse, memory blackouts, and concern expressed by others about the participant’s drinking, were reported at similar levels across the two groups.

### Alcohol-Related Consequences Among Injury Patients

Table 5 presents the distribution of alcohol-related consequences across the five DrInC subscales, comparing adults and young people. This analysis includes 645 current drinkers (98 young people and 547 adults) who reported alcohol use in the past 12 months. A statistically significant difference was observed in the physical subscale (*p* = 0.010), with a higher proportion of young people (85.7%) reporting alcohol-related physical consequences compared to adults (73.5%).

**Table 5:**
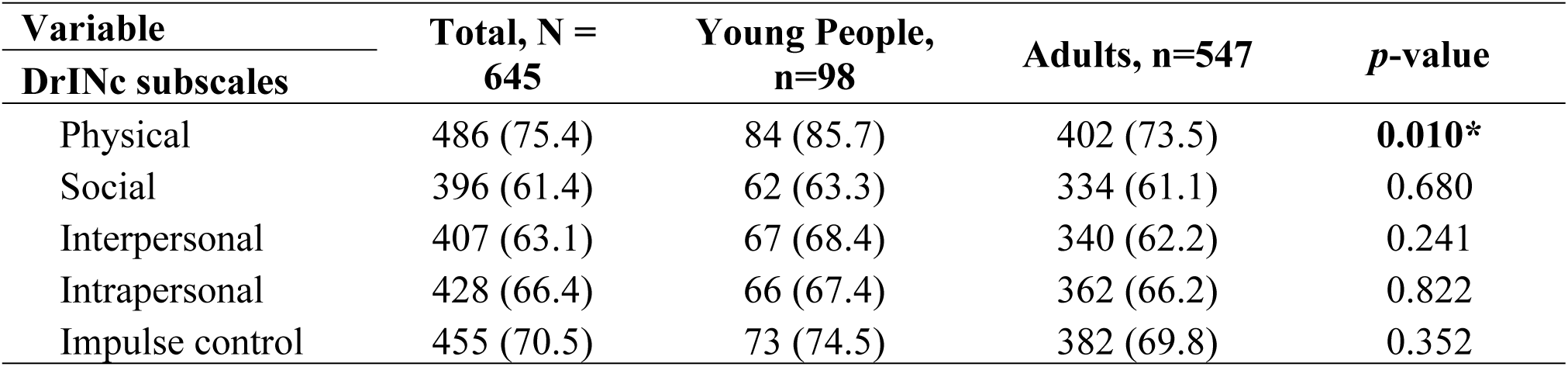
Comparison of Alcohol-Related Consequences across DrInC Subscales among Young People and Adults (N=645)

| Variable<br>DrInC subscales | Total, N =<br>645 | Young People,<br>n=98 | Adults, n=547 | *p*-value |
| --- | --- | --- | --- | --- |
| Physical | 486 (75.4) | 84 (85.7) | 402 (73.5) | **0.010\*** |
| Social | 396 (61.4) | 62 (63.3) | 334 (61.1) | 0.680 |
| Interpersonal | 407 (63.1) | 67 (68.4) | 340 (62.2) | 0.241 |
| Intrapersonal | 428 (66.4) | 66 (67.4) | 362 (66.2) | 0.822 |
| Impulse control | 455 (70.5) | 73 (74.5) | 382 (69.8) | 0.352 |

For the social subscale, 63.3% of young people and 61.1% of adults reported alcohol-related consequences, with no statistically significant difference between the groups (p = 0.680). Similarly, in the interpersonal subscale, 68.4% of young people and 62.2% of adults reported alcohol-related consequences (*p* = 0.241). The intrapersonal subscale showed comparable proportions of alcohol-related consequences among young people (67.4%) and adults (66.2%), with no statistically significant difference (p = 0.822). Likewise, for the impulse control subscale, 74.5% of young people and 69.8% of adults reported consequences, with no significant difference between groups (p = 0.352). (Table **5**)

## Discussion

To our knowledge, this is the first study in Tanzania to compare alcohol use patterns, Alcohol Use Disorder (AUD) screening by the AUDIT test, and alcohol-related consequences between young people and adults presenting with injuries. The findings highlight three key insights: First, while adults were more likely to report frequent alcohol consumption, young people were significantly more likely to report that they or someone else had been injured as a result of their drinking within the past year. Second, both age groups showed a similarly high prevalence of screening high for AUD, with no statistically significant difference. Third, young people reported significantly more physical consequences of alcohol use, suggesting that even with lower consumption frequency compared to adults, their drinking behavior is more associated with acute harms.

Young people were significantly more likely to report that they or someone else had been injured as a result of their drinking within the past year, despite drinking less frequently than adults. This finding is consistent with existing literature. Studies have found that younger drinkers are more susceptible to acute alcohol-related consequences due to a combination of lower alcohol tolerance, developmental vulnerability, and riskier drinking environments (25). For example, Hingson et al. (2017) (26) and Waller et al. (2018) (10) report that younger individuals are more likely to experience unintentional injuries and other harms following drinking. These patterns may be further shaped by misperceptions of drinking limits, poor coping mechanisms, and a lack of established harm-reduction strategies among younger people, which has also been documented in the Tanzanian context (26). Additionally, economic disparities may partly explain lower alcohol consumption among young people (27), as supported by the lower income observed in this group.

Despite the differences in drinking frequency and pattern, the comparable prevalence of AUD across young people and adults in this study indicates that harmful alcohol use is not confined to one developmental stage. Prior studies may have underemphasized more problematic drinking among people due to assumptions that younger individuals engage in riskier behaviors. Yet, recent research highlights increasing rates of AUD among adults (28), possibly due to shifting societal norms and reduced stigma, making older individuals more willing to disclose alcohol-related problems (29). These findings reinforce the importance of routine AUD screening in emergency settings regardless of age. However, alcohol-related harms may be more severe for young people, whose brains, particularly the prefrontal cortex, are still undergoing critical development. Thus, while AUD prevalence was similarly high across age groups, prevention and early intervention among young people are particularly urgent to mitigate lifelong consequences. School- and university-based alcohol education programs should emphasize the risks of early alcohol exposure, its impact on brain development, and the link to injury risk. Community outreach initiatives, including peer-led programs and digital campaigns, can further strengthen harm reduction messaging and promote safer behaviors.

Young people were also significantly more likely than adults to report physical consequences of alcohol use, as captured by the DrInC physical subscale. This includes immediate health outcomes such as vomiting, blackouts, and alcohol-related injuries, typical of early drinking experiences where tolerance is low and control strategies are not yet developed (30) . With age and prolonged exposure to alcohol, individuals may develop physiological tolerance or adapt behaviors that reduce acute harm, though this does not imply absence of risk. (31). Ibitoye et al. (2019) noted that adults in urban Tanzania often develop habitual consumption patterns that may escape immediate clinical attention but still contribute to long-term morbidity (32). Thus, while acute harms may be more visible in youth, chronic consequences must not be overlooked among adults.

### Limitations

The study has several limitations that should be acknowledged. First, the cross-sectional design limits our ability to establish causal relationships between alcohol use patterns and injury outcomes. Second, data on alcohol use and consequences were self-reported, introducing the possibility of recall bias and social desirability bias, particularly in a clinical setting. Although validated tools such as the AUDIT and DrInC were used, underreporting or misclassification of alcohol use may still have occurred. Third, the sample includes only injury patients who were clinically sober and consented to participate, potentially excluding those with more severe alcohol intoxication or cognitive impairments, thereby underestimating the true burden of alcohol-related harm. Finally, although the findings are based on a single referral hospital in Northern Tanzania and may not be generalizable to all settings, the hospital serves a wide catchment area encompassing both urban and rural populations, suggesting that the data are reasonably reflective of diverse patient experiences in the region.

### Conclusion

This study highlights important age-related differences in alcohol use patterns and alcohol-related harms among injury patients in Northern Tanzania. While adults reported more frequent alcohol consumption, young adults experienced disproportionately higher alcohol-related physical consequences and were more likely to report a recent instance, within the past year, where they or someone else had been injured as a result of their drinking. Despite these behavioral distinctions, both groups showed similarly high prevalence of alcohol use disorder, underscoring the widespread nature of harmful drinking in this population. These findings emphasize the need for age-tailored interventions within emergency care and public health systems, particularly strategies that address risky drinking behaviors among young adults and sustained use among older adults. Future research should explore the effectiveness of such targeted interventions, especially in low-resource settings, to reduce the burden of alcohol-related injuries and long-term health consequences.

## List of Abbreviations

AUD: Alcohol Use Disorder
AUDIT: Alcohol Use Disorders Identification Test
BAC: Blood Alcohol Concentration
DALY: Disability-Adjusted Life Year
DrInC: Drinker Inventory of Consequences
ED: Emergency Department
EMD: Emergency Medicine Department
HIC: High Income Countries
IQR: Interquartile Range
KCMC: Kilimanjaro Christian Medical Center
LMICs: Low- and Middle-Income Countries
PHQ-9: Patient Health Questionnaire-9
PRACT: Pragmatic Randomized Adaptive Clinical Trial
SD: Standard Deviation
SSA: Sub-Saharan Africa
TLFB: Timeline Followback
TZS: Tanzanian Shilling
WHO: World Health Organization

## Declarations

### Ethics approval and Consent to participate

Ethical approval for this study was obtained from the Kilimanjaro Christian Medical University College Ethics Committee (Certificate #2457) and the Tanzanian National Institute for Medical Research (NIMR/HQ/R.8a/Vol.IX/3425). Consent to participate was not required for this analysis, as it involved secondary use of de-identified data.

### Consent for publication

Not applicable

### Data Availability

Data used in this study is available upon reasonable request. Access to the dataset requires a written agreement approved by the Kilimanjaro Christian Medical Centre Ethics Committee and the National Institute for Medical Research (Tanzania)

### Competing Interest

The authors declare that they have no competing interest

### Funding

This study was supported by the Trauma Research Capacity Building Program at KCMC, funded through the D43 TRECK grant (#D43-TW012205) from the U.S. National Institutes of Health (NIH) and the Duke Department of Emergency Medicine (principal investigators: Mmbaga and Staton). WM and BM received support through the NIH-funded TRECK D43 program. EMK is funded by the Eunice Kennedy Shriver National Institute of Child Health and Human Development (grant number K23 HD112548). The PRACT Trial was supported by the National Institute on Alcohol Abuse and Alcoholism (NIAAA) of the NIH, grant number 1R01AA027512-01A1 (principal investigator: Staton). The funders had no role in study design, data collection, analysis, interpretation, or manuscript preparation.

### Author Contribution

WM, CS, SEM, JV, FJM and BTM were involved in conceptualizing the study and formulating the research question and rationale. WM led the data analysis and drafted the initial manuscript. EMK, BM, AA, BTM, RNF, CS, SEM, FJM, and JV contributed to manuscript review, interpretation of findings, and critical revisions. All authors reviewed and approved the final manuscript.

## Acknowledgement

The authors acknowledge the contributions of the research team at the Global Emergency Medicine Innovation and Implementation (GEMINI) Research Center, the KCMC-Duke Collaboration, and the Institute of Public Health at Kilimanjaro Christian Medical University College. Special thanks to the study participants who made this study possible.

## Notes

### Competing Interest Statement

The authors have declared no competing interest.

